# Dyspareunia in their own words: A comprehensive qualitative description of endometriosis-associated sexual pain

**DOI:** 10.1101/19005793

**Authors:** Kate J Wahl, Shermeen Imtiaz, KS Joseph, Kelly B Smith, Paul J. Yong, Susan M. Cox

## Abstract

**Background:** Dyspareunia is a classic symptom of endometriosis but is neglected in research and clinical contexts. This study explored the experience of this endometriosis-associated sexual pain.

**Methods:** This was a qualitative descriptive study that included people who had experienced endometriosis-associated dyspareunia alone or with a partner. Data collection involved semi-structured interviews with a female researcher that began with an open-ended question about dyspareunia and included interview prompts related to the nature of sexual pain. Interviews were recorded, transcribed verbatim, and analysed for themes.

**Results:** 17 participants completed interviews. The mean participant age was 33.3 (SD=7.2) and most participants identified as white (82%), were college-educated (71%), identified as heterosexual (65%), and were partnered (59%). Location, onset, and character emerged as important, interrelated features of endometriosis-associated dyspareunia, as did severity and impact. Dyspareunia occurred at the vaginal opening (n=7) and in the abdomen/pelvis (n=13). Pain at the vaginal opening began with initial penetration and had pulling, burning and stinging qualities. Pain in the pelvis was typically experienced with deep penetration or in certain position and was described as sharp, stabbing and/or cramping. Dyspareunia ranged from mild to severe, and for some participants had a marked psychosocial impact.

**Conclusions:** Dyspareunia is a heterogeneous symptom of endometriosis that ranges in severity and impact. Disaggregating dyspareunia into superficial and deep types may better reflect the etiologies of this pain, thereby improving outcome measurement in intervention studies and clinical care.

## INTRODUCTION

Endometriosis is a gynecological condition that affects 10 percent of females and is characterized by the growth of ectopic endometrial-type tissue. [1] More than half of people with endometriosis experience pain during sexual intercourse.[2,3] This pain, called dyspareunia, is associated with psychosocial sequalae including poor quality of life,[3,4] low self-esteem,[5–7] and relationship difficulties.[5,7,8]

Despite its prevalence and impact, dyspareunia is considered a ‘neglected symptom’ of endometriosis.[9] Clinically, its neglect has been attributed to embarrassment and normalization by clinicians and patients as well as a dearth of evidenced-based treatment options.[8,9] Dyspareunia has also been neglected in the research context, where it is not validly operationalized and studies are underpowered to detect relevant outcomes.[10]

Several qualitative studies have highlighted the impact of dyspareunia on the lives of people with endometriosis.[5–8,11,12] Qualitative data about dyspareunia have also been collected for the development of patient-reported outcome measures of endometriosis pain.[13–18] However, reporting of the results has not focused on sexual pain and demographic characterization of participants is often limited.

Largely absent from the literature is a complete description of what endometriosis-associated sexual pain feels like or where and when it happens. The aim of this study was therefore to provide a comprehensive account of dyspareunia in the everyday terms people with endometriosis use to describe the pain. Such data are required to support more accurate phenotyping of this symptom in clinical and research contexts.

## MATERIALS AND METHODS

### Design

This was a qualitative descriptive study. Compared to other approaches, qualitative description requires less interpretation and instead focuses on identifying and reporting the fundamental elements of an experience in the words of participants.[19]

### Patient and Public Involvement

The research conducted by our team is guided by a Patient Research Advisory Board. The Board approved the study design and was not involved with recruitment. Participants will receive the peer-reviewed article with a plain language summary of the results.

### Participants and Recruitment

Participants were recruited from the Endometriosis Pelvic Pain Interdisciplinary Cohort data registry (ClinicalTrials.gov Identifier: NCT02911090). The data registry is open to patients attending a tertiary centre for endometriosis and captures longitudinal demographic and clinical data. Potential participants were enrolled in the data registry and consented to be contacted for research. Inclusion criteria were age of 18 years or older, clinically suspected or diagnosed endometriosis, and current or previous dyspareunia alone or with a partner. Exclusion criteria were never sexually active alone or with a partner, no history of dyspareunia alone or with a partner, and inability to communicate in English. Recruitment continued until saturation, the point when additional interviews did not reveal new insights about the experience of sexual pain.

To reflect the demographic characteristics of patients seen at the tertiary centre, potential participants were randomly selected from within overlapping education, sexual orientation, and ethnicity groups. KW contacted selected participants by phone to present the study, introduce herself and her program of research, and screen for eligibility. Interested participants received and returned consent forms by e-mail. To reduce bias, each selected participant was called a minimum of five times and at different times of day before a subsequent participant was selected.

### Data Collection and Analysis

The interview guide was developed by KW, a graduate student with theoretical and applied training in qualitative methodologies, SC a qualitative health researcher focused on illness experiences throughout the life course, PY a clinician-scientist with expertise in endometriosis, and members of the Patient Research Advisory Board for the Centre for Pelvic Pain and Endometriosis. KW pilot tested the interview guide and conducted a single one-on-one semi-structured telephone interview with each participant. Each interview began with the question “Tell me about the pain you experience with sex”; initial prompts related to *a priori* themes of pain site, onset, character, radiation, associations, time course, and exacerbating/relieving factors. Contemporaneous field notes permitted the identification of emergent themes and assessment of saturation. Demographic and clinical data were drawn from the data registry.

The interviews were audio recorded and transcribed verbatim by KW, SI, and a research assistant. Using Nvivo 12 Pro,[20] KW conducted a qualitative content analysis of the transcripts.[21] The analysis involved reading the set of transcripts to get a global sense of the data; reading transcripts individually and highlighting words that captured key concepts as codes; sorting the codes into categories based on relatedness; and organizing the categories into meaningful clusters.

To ensure reliability of the analysis, SI coded two randomly selected transcripts and cross-checked her findings with those of KW. Transcript and analysis checking with the participants was not completed given the low-inference approach to analysis.

## RESULTS

### Participants

Of 50 potential participants we attempted to contact about the study, 17 completed an interview, 11 could not be reached, 10 were ineligible, 4 declined to participate, and 8 were lost to follow up. The average age of the participants was 33.3 years (SD=7.2, range 23-50). Race, gender identity, sexual orientation, educational attainment, marital status, parity, and endometriosis status are described in Table 1. Mean dyspareunia scores for initial penetration and deep penetration on an 11-point numeric rating scale (0=no pain, 10=worst pain imaginable) were 3.3 (SD=3.4, range 0-8) and 6.6 (SD=2.8, range 0-10) respectively. The average interview length was 28 minutes (SD=13, range 14-72).

**Table 1.** Demographic characteristics of the study sample

| **Characteristic** | **mean ± SD,**<br>**range; n (%)** |
| --- | --- |
| Age | 33.5 ± 7.0, 23-50 |
| Race |  |
| White | 14 (82) |
| Asian | 2 (12) |
| Other | 1 (6) |
| Female Gender Identity | 17 (100) |
| Sexual Orientation |  |
| Heterosexual | 12 (70) |
| Bisexual | 2 (12) |
| Lesbian | 2 (12) |
| Other | 1 (6) |
| Education |  |
| High school (grades 9-12) | 1 (6) |
| Community college/vocational school | 3 (18) |
| College | 9 (52) |
| Graduate school | 4 (24) |
| Marital Status |  |
| Single | 2 (12) |
| Dating | 2 (12) |
| Common law | 2 (12) |
| Married | 9 (52) |
| Separated | 2 (12) |
| Parous | 4 (24) |
| Endometriosis status |  |
| Clinically suspected | 10 (59) |
| Surgical (visual) diagnosis | 1 (6) |
| Surgical (histological) diagnosis | 7 (41) |
| Current endometrioma on ultrasound | 2 (12) |
| Current nodule on examination | 2 (12) |
| Deep dyspareunia score | 6.76 ± 2.82, 0-10 |
| Superficial dyspareunia score | 3.24 ± 3.33, 0-8 |

Although five participants did not self-identify as heterosexual in the data registry, only one explicitly mentioned her sexual orientation in the interview. These participants did not report experiences of sexual pain different from those of other participants.

### Pain Fundamentals

Table 2 shows that across participants, pain began at different points in the sexual encounter. The table also shows that participants used a range of terms to describe the character and anatomic location of their endometriosis-associated sexual pain.

**Table 2.**
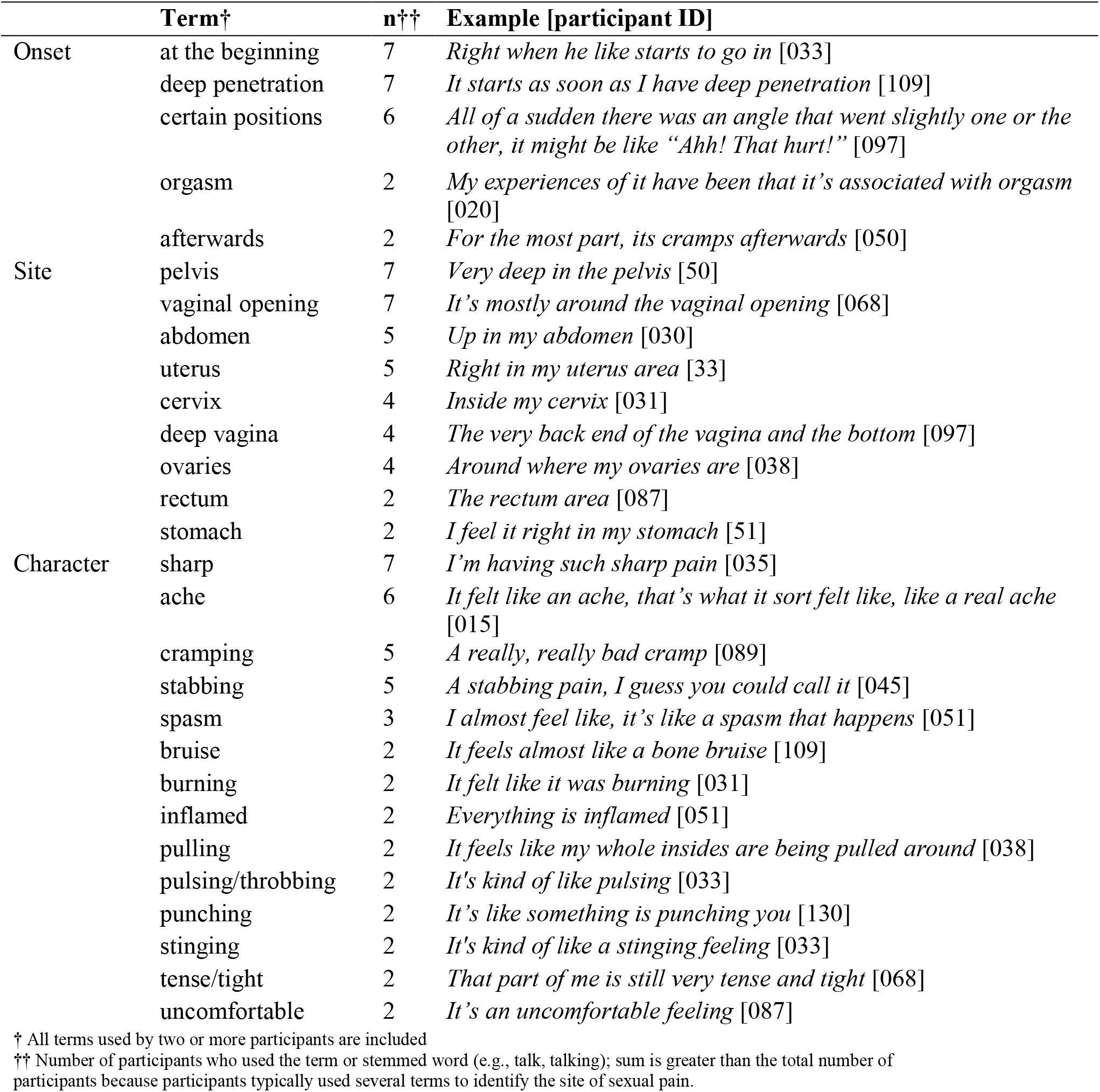
Terms used to describe sexual pain

Some participants had difficulty naming the site of their pain and occasionally explained its location in terms of other painful experiences.

> *I’m not really not familiar with my biology, the reproductive parts. But the inside. So is it the cervix? Or the uterus?* [130]
>
> *That’s really hard to explain, because I can’t really pinpoint where it is. It’s not on the outside, it’s the inside where, if I was pointing at my pelvis, like a couple inches in*. [089]
>
> *The same place I would experience period cramps*. [051]

All participants experienced pain in the pelvis or pelvic organs and seven participants also experienced pain at the vaginal introitus. For example, participant 020 said,

> *I have a few different things with pain with penetration*.

This participant went on to clarify,

> *So I experience the three types of penetrative pain so like the penetrative pain in my vagina, and then deep vaginal pain, and I also experience uterine pain with arousal and orgasm*.

Except for two participants who reported pelvic pain at orgasm and two who reported pelvic pain after intercourse, pain in this area began with deep penetration or certain sexual positions and was associated with sharp, aching, cramping or stabbing feelings. In contrast, pain at the vaginal opening always began with initial penetration and was associated with pulling, stinging, or burning.

### Pain Severity

When asked about the severity of their pain, 14 participants spontaneously rated their pain as a score out of 10. Participants also described the severity of their pain.

> *It’s not so bad where you’re like heeled over, like when you have your period with endo, where you can’t move or anything like that, it’s not even close to that, it’s just, it’s more uncomfortable I’d say*. [030]
>
> *I could keep going if I mentally prepared myself but there is also like a side to me that would just want to stop*. [033]
>
> *So, it wasn’t the worst pain I’d experienced but it was uncomfortable enough that I would have to stop doing that activity*. [043]
>
> *It wasn’t quite at a level I would pass out from the pain, but I would definitely cry quite a bi*t. [031]
>
> *The worst pain I’ve ever felt. Nothing could be done, I can’t have people around me, I don’t like noises, I just want everyone to just leave me alone. There’s nothing that can be done for it to really go away you know, until it goes away. I don’t know, it’s scary*. [051]

### Pain Impact

Fifteen participants reported having interrupted sex because of pain and ten had avoided sexual experiences.

The experience of sexual pain affected the emotional and psychological well-being of some participants.

> *It’s hard for me to just deny all the time because I start to feel bad*. [033]
>
> *It would make me feel sort of you know, emotionally kind of discouraged*. [043]
>
> *I feel insignificant, you almost feel broken or something*. [051]
>
> *You feel guilty for having to stop*. [089]
>
> Many participants also reported that the experience of sexual pain impacted their intimate relationships.
>
> *I think my husband was reluctant to approach me for sex because he knew I was feeling ill. You know, you don’t want to ask somebody who’s feeling ill to “Hey let’s go!” I’m glad we’ve got a solid marriage and we’ve weathered the storms and we’re ok now. But it was a really tough time*. [043]
>
> *I don’t ever want to have sex, I would be happy to not do it at all, but I have to get myself in the mood. I try once a month to have, you know, that closeness with him even though for me its not pleasurable at all […]. It has kept me keeping some secrets from my lover, which is not how our relationship normally is*. [038]
>
> *Lately, we just have sex is because we want to conceive, or try to conceive [*…*] we don’t really have sex just because we wanted to*. [130]
>
> *It got to the point where I was in pain so often from intercourse, that my partner and I basically decided that we just weren’t going to anymore, that it just wasn’t worth me going through the pain […] It contributed to the end of that relationship*. [031]
>
> For one participant who was single, sexual pain affected her willingness to seek out physical intimacy.
>
> *I don’t want to watch the horrified faces as I’m suddenly leaping out of bed because I’m in so much pain*. [035]
>
> Some participants had sexual intercourse infrequently or not at all. Four participants reported that hormonal treatment relieved their pain, but in one instance the treatment also negatively affected sexual desire. Other participants found ways of managing and coping with their pain.
>
> *I typically have to take anti-inflammatories for the pain and just managing it, or a hot water bottle, a hot shower, that sort of thing*. [109]
>
> *We went out and bought that masturbation sleeve and we kind of control the depth just to kind of prevent [the pain] from happening*. [051]
>
> *I just think about noises or him speaking to me and I really just focus on that. I take my mind away from what’s actually happening*. [033]
>
> *I have pain with penetration almost all the time, but I am able to accommodate it in a way so it’s not severe enough that it stops the experience and like I can change a position or stop having penetration and it’s fine and I can continue with my sexual experience*. [020]
>
> *I’ve gone to therapy to try to stop myself from feeling guilty right away and just be okay with the fact that like, we might have to change things up and that’s okay*. [089]

## DISCUSSION

This study set out to identify the fundamental features of endometriosis-associated dyspareunia. Participants reported pain in the pelvis or pelvic organs, which generally began with deep penetration or certain positions and had a sharp, aching, cramping, or stabbing quality. Almost half of participants also experienced pain at the vaginal opening that began with initial penetration and had a pulling, burning, or stinging nature. This pain ranged in severity and in some cases had a significant, negative impact.

Although participants sometimes struggled to describe the location of their sexual pain, our study showed that dyspareunia typically occurred in the pelvis and at the vaginal opening. These results aligned with research by Fauconnier et al. that identified five themes associated with sexual pain: strong and sharp pain, deep internal pain, pain in certain positions, distracting pain that prevents or disrupts intercourse, and a burning feeling during or after intercourse.[22] Clinically, the finding of pelvic pain with deep penetration corresponds with the understanding of endometriosis-associated dyspareunia whereas the description of pain at the vaginal opening that begins with initial penetration mirrors the presentation of other gynecological conditions like provoked vestibulodynia.[23]

Important secondary findings of this study related to the impact and management of endometriosis-associated sexual pain. Dyspareunia had a variety physical, emotional, mental, and interpersonal effects that participants managed with medical, physical, and cognitive interventions. These results aligned with previous work recommending a biopsychosocial model and interdisciplinary treatment approach for endometriosis-associated sexual pain.[24,25]

The defining strength of the study was its exclusive and in-depth approach to characterizing endometriosis-associated sexual pain. The findings confirmed and extended those of Fauconnier et al., who described dyspareunia using data from 10-minute French-language interviews about painful symptoms of endometriosis.[22] Additional strengths were that the study sample was selected to reflect the demographic characteristics of the clinical population and that participants ranged in severity of their self-reported sexual pain.

An obvious limitation of the study was that participants were not selected based on gold-standard surgical diagnosis (Table 2). However, this reflects a shift towards clinical diagnosis in practice.[26] A second limitation was that all participants had previously attended a tertiary centre where they completed dyspareunia measures and discussed the sexual pain with a care provider. Together with high levels of education in the study sample, earlier clinical interactions may have affected how participants described their pain.

Our findings have implications for endometriosis-associated dyspareunia in clinical research. First, measuring sexual pain without distinguishing between pelvic pain and pain at the vaginal opening may capture dyspareunia that arises from other conditions,[27] thereby leading to misclassification and potentially biasing the effect of interventions toward the null. Second, reliance on onset- or location-based vocabulary in patient-reported measures might limit response accuracy; diagrams that highlight the relevant anatomic sites could facilitate true responses. Third, although recent clinical trials have used ordinal response options (i.e., none, mild, moderate, severe),[28] reporting dyspareunia on a scale of 10 is intuitive for respondents and aligns with recommendations for the measurement of patient-important endometriosis pain symptoms.[29] Finally, research focused on sexual pain should seek to account for the physical and emotional impact of this symptom.

Clinically, our findings suggested that people who experience endometriosis-associated dyspareunia have unmet treatment needs. Asking whether a patient has any concerns regarding pain with sexual activity that they would like to discuss can avoid neglect of this symptom due to taboos or normalization.[5,6] For patients who consider their sexual pain a priority, determining whether the pain occurs in the pelvis or at the vaginal opening will help guide additional investigations and treatments. Involving intimate partners in counselling about dyspareunia with the consent of both parties can also improve the patient-centeredness of care.[30]

Dyspareunia is a primary symptom of endometriosis, yet has been relegated to a secondary or tertiary outcome in clinical trials research.[28,29] Other studies have established that endometriosis-associated sexual pain negatively affects physical and psychosocial well-being, a finding that the present work reinforced. This study also provided a comprehensive insight into the experience of dyspareunia in endometriosis, yielding methodological considerations for how this symptom should be measured as well as clinical implications. We hope that by providing a rich description of endometriosis-associated sexual pain, this work will be hypothesis-generating and will contribute to rigorous investigation of dyspareunia in endometriosis.

## Data Availability

No data are available

